# The removal of airborne SARS-CoV-2 and other microbial bioaerosols by air filtration on COVID-19 surge units

**DOI:** 10.1101/2021.09.16.21263684

**Authors:** Andrew Conway-Morris, Katherine Sharrocks, Rachel Bousfield, Leanne Kermack, Mailis Maes, Ellen Higginson, Sally Forrest, Joana Pereira-Dias, Claire Cormie, Tim Old, Sophie Brooks, Islam Hamed, Alicia Koenig, Andrew Turner, Paul White, R. Andres Floto, Gordon Dougan, Effrossyni Gkrania-Klotsas, Theodore Gouliouris, Stephen Baker, Vilas Navapurkar

## Abstract

**Background:** The COVID-19 pandemic has overwhelmed the respiratory isolation capacity in hospitals; many wards lacking high-frequency air changes have been repurposed for managing patients infected with SARS-CoV-2 requiring either standard or intensive care. Hospital-acquired COVID-19 is a recognised problem amongst both patients and staff, with growing evidence for the relevance of airborne transmission. This study examined the effect of air filtration and ultra-violet (UV) light sterilisation on detectable airborne SARS-CoV-2 and other microbial bioaerosols.

**Methods:** We conducted a crossover study of portable air filtration and sterilisation devices in a repurposed ‘surge’ COVID ward and ‘surge’ ICU. National Institute for Occupational Safety and Health (NIOSH) cyclonic aerosol samplers and PCR assays were used to detect the presence of airborne SARS-CoV-2 and other microbial bioaerosol with and without air/UV filtration.

**Results:** Airborne SARS-CoV-2 was detected in the ward on all five days before activation of air/UV filtration, but on none of the five days when the air/UV filter was operational; SARS-CoV-2 was again detected on four out of five days when the filter was off. Airborne SARS-CoV-2 was infrequently detected in the ICU. Filtration significantly reduced the burden of other microbial bioaerosols in both the ward (48 pathogens detected before filtration, two after, *p*=0.05) and the ICU (45 pathogens detected before filtration, five after *p*=0.05).

**Conclusions:** These data demonstrate the feasibility of removing SARS-CoV-2 from the air of repurposed ‘surge’ wards and suggest that air filtration devices may help reduce the risk of hospital-acquired SARS-CoV-2.

**Funding:** Wellcome Trust, MRC, NIHR

## Introduction

During the COVID-19 pandemic ‘general’ hospital wards in the UK were rapidly repurposed into ‘surge’ wards and intensive care units (ICU), which lacked the capacity for high frequency air-changes. Airborne dissemination is likely an important transmission route for SARS-CoV-2 ^1^, with SARS-CoV-2 RNA being detected in air samples from wards managing COVID-19 patients ^2,3^. Despite the use of appropriate personal protective equipment (PPE) that filter medium and large size droplets, there are multiple reports of patient-to-healthcare worker transmission of SARS-CoV-2 ^4,5,6,^ potentially through the inhalation of viral particles in small (< 5µM) aerosols ^7^. Furthermore, nosocomial acquisition of COVID-19 has continued to blight healthcare systems despite the systematic introduction of patient and healthcare worker asymptomatic screening programmes^8^. There is a need to improve the safety for healthcare workers and patients during the pandemic by decreasing the potential for the airborne transmission of SARS-CoV-2^7^. Engineering solutions that improve ventilation with provision of UV light sterilisation are considered a more effective intervention in the hierarchy of controls against transmissible infections compared to enhanced respiratory protective equipment^9,10^. Portable air filtration systems, that combine high efficiency particulate filtration and ultraviolet (UV) light sterilisation, may be a scalable solution for removing respirable SARS-CoV-2. A recent review by the UK Scientific Advisory Group for Emergencies modelling group found limited data regarding the effectiveness of such devices^11^, which is consistent with findings from two recent systematic reviews ^12, 13^. Most of the testing of such systems has been physical device validation using inorganic particles or removal of bacterial bioaerosols in controlled test environments ^12,13^. Here we present the first data providing evidence for the removal of SARS-CoV-2 and microbial bioaerosols from the air using portable air filters with UV sterilisation on a COVID-19 ‘surge’ ward during the ongoing pandemic.

## Methods

### Setting

The study was conducted in two repurposed COVID-19 units in Addenbrooke’s Hospital, Cambridge, UK in January/February 2021 when the alpha variant (lineage B1.1.7) comprised >80% of circulating SARS-CoV-2^8^. One area was a ‘surge ward’ (ward) managing patients requiring simple oxygen therapy or no respiratory support, the second was a ‘surge ICU’ (ICU) managing patients requiring invasive and non-invasive respiratory support. The ward was a fully occupied four-bedded bay (Fig. 1A). The ICU was fully occupied five-bedded bay, with a super-capacity sixth occupied bed used in week 2 (Fig. 1B).

**Figure 1.**
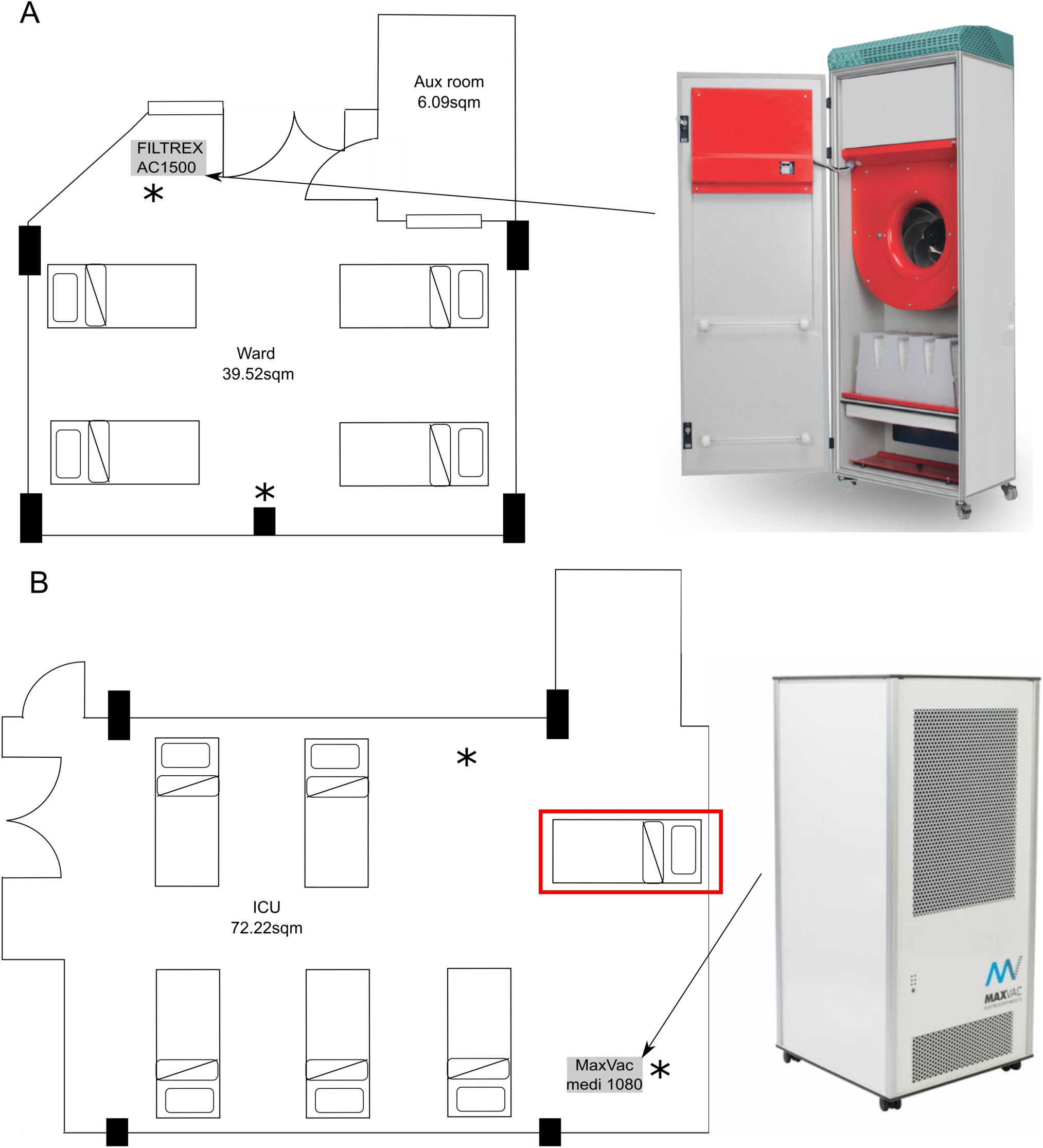
Location of the air filters and room layout. A) Layout of the room on the ‘surge’ ward with four beds. B) Layout on the ‘surge’ ICU with six beds including the addition of the additional bed to increase occupancy (labelled with rad box). Locations of the NIOSH air samplers indicated by *. The air filters were installed in the marked locations and set to operate at 1000 m^3^/hour. The rooms volumes are approximately 107 m^3^ and 195m^3^ respectively. Fresh air was not supplied or extracted in these areas.

In the ward we installed an AC1500 HEPA14/UV steriliser (Filtrex, Harlow, UK), whilst in the ICU we installed a Medi 10 HEPA13/UV steriliser (Max Vac, Zurich, Switzerland) (supplemental methods). The air filters were placed in fixed positions before the initiation of the three-week study period (Fig. 1), switched on at the beginning of week two and run continuously from Sunday to Sunday for 24 hours per day, providing approximately 5-10 room-volume filtrations per hour in each location. As the devices do not meet medical device electrical safety standards (EN60601) they were operated at a distance of ≥1.5metres from any patient.

### Study design

We performed a crossover evaluation, with the primary outcome being detection of SARS-CoV-2 RNA in the various size fractions of the air samples. Air sampling was conducted using National Institute for Occupational Safety and Health (NIOSH) BC 251 two-stage cyclone aerosol samplers^12^ (Donated by B Lindsley, Centers for Disease Control, Atlanta), operated in accordance with previous studies demonstrating capture of airborne viruses (supplemental methods)^2,14-18^. Air samplers were assembled daily with a sampler left in a sealed bag as a control. Samplers placed adjacent to the air filter inlet and the other at approximately four meters and no closer than two meters to patients (Fig. 1). In ICU two distant samplers were used, one mounted at head height and one at bed height.

The samplers were operated on weekdays (0815hrs to 1415hrs) for three consecutive weeks. After sampling, the samplers were disassembled using sterile technique and the filter papers were transferred to 15 ml Falcon tubes. The samples were processed then stored at −80°C until analysis. The samplers were washed with 80% ethanol and demineralised water.

### Pathogen detection

Nucleic acids were extracted from each NIOSH sampler component (tubes containing large aerosols, medium aerosols, and filter), as previously described^19^. Details of the RT-qPCR for SARS-CoV-2 and multiplex qPCR assays for a range of respiratory and other bacterial, viral, and fungal pathogens are in the supplemental section.

### Statistical analyses

Differences in the number of pathogens detected when air filter was on and off were compared by Mann-Whitney U-test. Statistical significance was inferred when *p* values were ≤0.05. Graphs were generated in R studio.

The study was registered as a service evaluation with Cambridge University Hospitals NHS Foundation Trust (Service Evaluation Number PRN 9798).

## Results

### Removal of SARS-CoV-2 by air filtration on surge ward

For the duration of the study (18^th^ January to 5^th^ February) the beds in the ward and ICU were at 100% occupancy, with 15 patients admitted to the ward and 14 admitted to the ICU over the three-week sampling period (7, 4, 4 in weeks 1-3 in the ward and 6, 5, 3 in the ICU, respectively). All patients were symptomatic and tested positive for SARS-CoV-2 RNA from a respiratory sample before admission. Patients in the ICU were managed with non-invasive mask ventilation, high flow nasal oxygen or invasive ventilation via endotracheal tube or tracheostomy. Patients in the ward were spontaneously ventilating with simple oxygen therapy or no respiratory support and no aerosol-generating procedures performed.

In the ward, during the first week whilst the air filter was inactive, we were able to detect SARS-CoV-2 on all five sampling days; RNA was detected in both the medium (1-4μM particle size) and the large (>4μM particle size) particulate fractions (Fig. 2A). SARS-CoV-2 RNA was not detected in the small (<1μM) particulate filter. The air filter was switched on in week two and run continuously; we were unable to detect SARS-CoV-2 RNA in any of the sampling fractions on any of the five testing days. These initial observations provided evidence for the removal of SARS-CoV-2 via the air filter system, albeit at high baseline C_*T*_ values. To confirm this observation, we completed the study by repeating the sampling with an inactive air filter. As in week one, we were able to detect SARS-CoV-2 RNA in the medium and the large particulate fractions on 3/5 days of sampling (a sample without tube size indicated tested positive on day 5) (Fig. 2A). We did not detect SARS-CoV-2 RNA from the control sampler.

**Figure 2.**
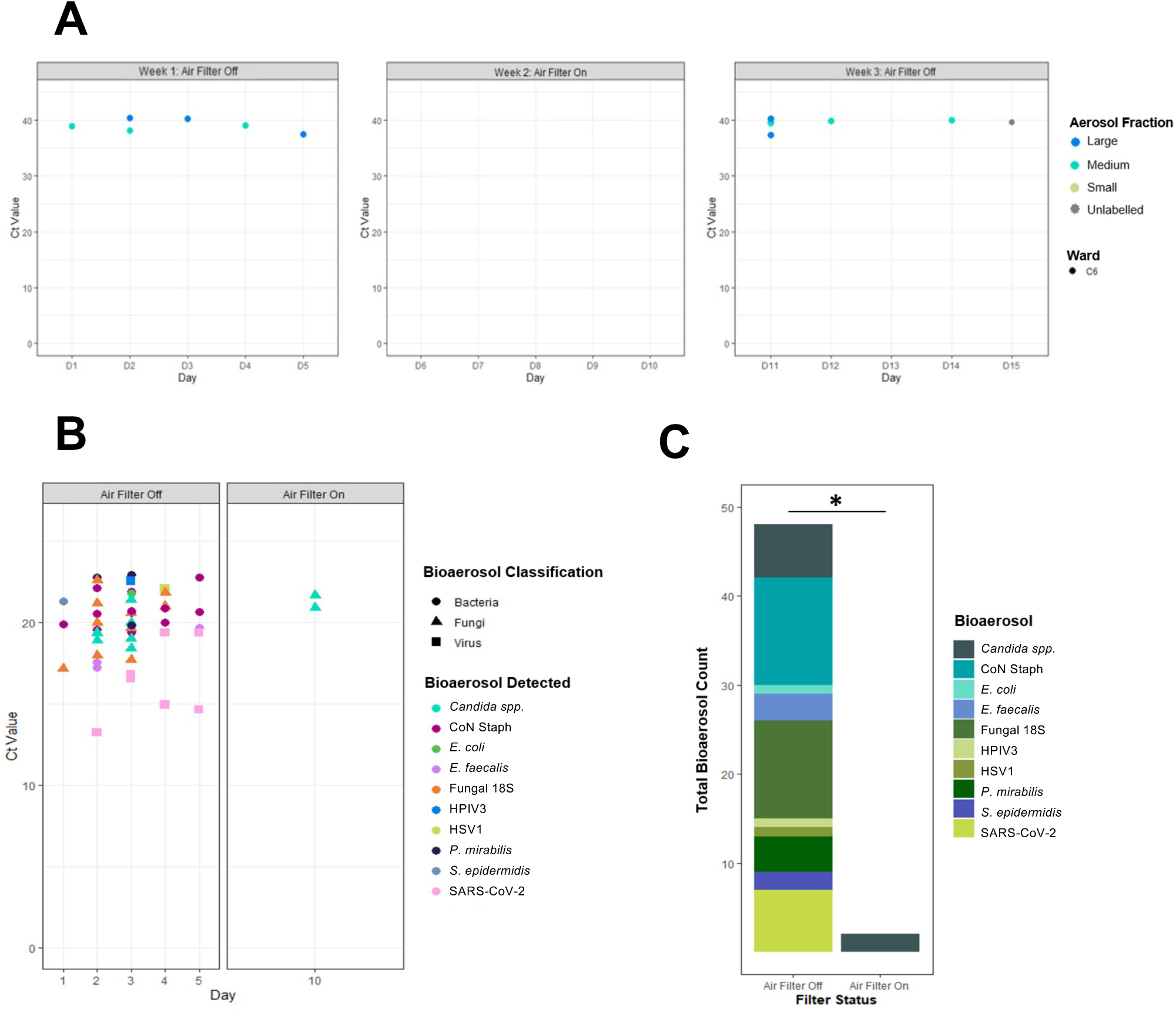
Bioaerosol detection in specific air sampler fractions over the three-week testing period on the ‘surge’ ward. A) C_*T*_ values for SARS-CoV-2 qPCR on air sample fractions collected daily from the ward. Colours indicate the specific component of the sampler where SARS-CoV-2 was detected. Components collected aerosols dependent on size fractions; large >4 µm, medium1-4 µm, small <1 µm. B) Daily detection of fungal, bacterial and viral bioaerosols detected by high-throughput qPCR collected during weeks one (filter off) and two (filter on). The differences in C_*T*_ values between the regular qPCR (A) and high-throughput qPCR (B) are a function of the microfluidics technology, and do not reflect higher bioaerosol burdens. C) Stacked bar chart showing collated total number of bioaerosol detections during weeks one (filter off) and two (filter on). \**p*=0.05 by Mann-Whitney U test.

### Removal of additional bioaerosols by air filtration on surge ward

We subjected the extracted nucleic acid preparations to high-throughput qPCR using a Biomark HD system to detect a range of viral, bacterial, and fungal targets. In the week one samples, we detected nucleic acid from multiple viral, bacterial, and fungal pathogens on all sampling days (Fig. 2B). In contrast, when the air filter was switched on, we detected yeast targets only on a single day, with a significant reduction (*p*=0.05) in microbial bioaerosols when the air filter was operational (Fig. 2C). Using this high-throughput approach, SARS-CoV-2 RNA was detected on 4/5 days tested in week 1 but was again absent in week 2. We were unable to generate multiplex data for week three due to sample degradation after storage of sample following SARS-CoV-2 RNA amplification.

### Effectiveness of air filter on surge ICU

In contrast to the ward, we found limited evidence of airborne SARS-CoV-2 in weeks one and three (filter off) but detected SARS-CoV-2 RNA in a single sample in the medium (1-4μM particle size) particulates on week 2 (filter on) (Fig. 3A). This contrary result did not reflect a general lack of bioaerosols in the ICU, which demonstrated a comparable quantity and array of pathogen associated nucleic acids to that seen in the unfiltered ward air on week one (Fig. 3B). Again, the use of the air filtration device significantly (*p*=0.05) reduced the microbial bioaerosols (Fig 3C); with only three organism types detected on two of the sampling days (Fig 3B). SARS-CoV-2 RNA was only detected once on the high-throughput qPCR assay, during week one.

**Figure 3.**
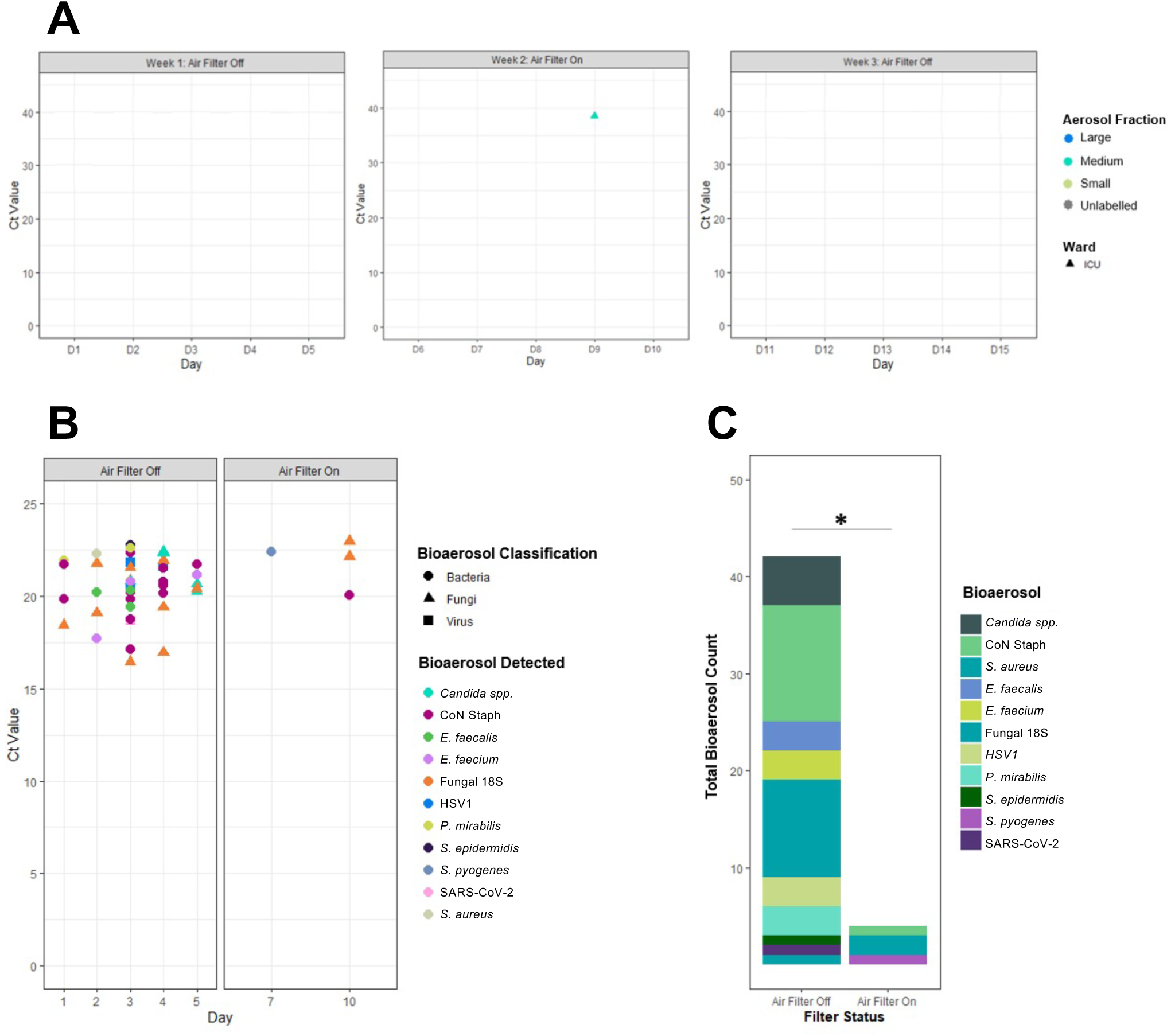
Bioaerosol detection in specific air sampler fractions over the three-week testing period on the ‘surge’ ICU. A) C_*T*_ value for the single qPCR SARS-CoV-2 detection on day 9 (week 2) in the medium (1-4 µm particle size) fraction. B) Daily detection of fungal, bacterial and viral bioaerosol detected by high-throughput qPCR collected during weeks one (filter off) and two (filter on). The differences in C_*T*_ values between the regular qPCR (A) and high-throughput qPCR (B) are a function of the microfluidics technology, and do not reflect higher bioaerosol burdens. C) Stacked bar chart showing collated total number of bioaerosol detections during weeks one (filter off) and two (filter on). \**p*=0.05 by Mann-Whitney U test.

## Discussion

Our study represents the first report of successful removal of airborne SARS-CoV-2 in a hospital environment using combined air filtration and UV sterilisation technology. Specifically, we provide evidence for the circulation of SARS-CoV-2 in a ward within airborne droplets of >1μM. Droplets of 1-4μM are likely a key vehicle for SARS-CoV-2 transmission, as they can remain airborne for a prolonged period. They are also readily respirable and can deposit in the distal airways. Recent data has shown that exertional respiratory activity, such as that seen in patients with COVID-19, increases the release of 1-4 μM respiratory aerosols, whilst conventionally defined ‘aerosol generating procedures’ such as high flow nasal oxygen and non-invasive ventilation actively reduce aerosol generation during exertion^20^. These data are consistent with our observations, suggesting that precautions to remove aerosolisation may be more important in conventional wards than in well defined ‘aerosol risk areas’. We also found a low burden of SARS-CoV-2 in the air on the ICU. This observation, combined with the higher level of aerosol protection worn by ICU staff, may explain why staff in these areas appear to be at significantly lower risk of acquiring COVID than those working on wards^21^.

The sampling and detection of airborne viruses poses several technological challenges, and although several approaches have been developed, there remains no agreed standard for their use or interpretation^22^. However, the detection of SARS-CoV-2 RNA by RT-qPCR (albeit at a high C_*T*_ value), and the lack of detection during use of an air filtration/UV sterilisation system, adds to a growing body of evidence implicating the airborne transmission of SARS-CoV-2 ^1^. The detection of SARS-CoV-2 RNA in the air of a ward managing patients with COVID-19 intimates that this is a key mechanism by which healthcare professionals could become infected during patient care. The removal of airborne viral particles and other pathogens may help reduce the likelihood of hospital-acquired respiratory infections. This reduction may be by both decreasing the load of respirable particles and by removal of larger droplets that can facilitate fomite-associated spread^22^. The clearance of bioaerosol was not restricted to SARS-CoV-2. A range of bacteria, yeasts, and other respiratory viruses with pathogenic potential were detected in the air of both rooms in the first week, with their burden significantly reduced during air filtration. Although the impact of air filtration on nosocomial infection is uncertain^23^, the broad range of pathogens removed in this study suggests potential for benefit beyond removal of SARS-CoV-2.

There are several potential explanations for the lower detection of SARS-CoV-2 in air of an ICU. These include a later stage of disease during which viral replication is less pronounced^24^, higher viral loads in the lower rather than upper respiratory tract in critically ill patients^25^ and use of respiratory devices, which reduce aerosol generation^20^. The reduction in microbial bioaerosols found in ICU during the week of the air filtration system provides confidence that the device was similarly effective to that used on the ward, despite the infrequent detection of SARS-CoV-2.

A recent systematic review of both static and portable air filtration, which also assessed relevant building codes and guidelines^12^, identified no robust studies of air filtration. Although multiple building codes propose air filtration to protect vulnerable patients and to reduce risks of transmission of airborne diseases, these have not been updated in light of COVID-19^12^. Mousvai and colleagues identified several studies demonstrating the capacity of air filtration to reduce inert, fungal, and bacterial bioaerosols in experimental and clinical contexts. These findings are consistent with our report, but previous data originate from fixed rather than portable air filtration devices. No reports of SARS-CoV-2 removal were identified. A further recent review focussed solely on portable air filters^13^, with studies demonstrating the removal of inert particles and deliberately aerosolised bacteria, again no reports of SARS-CoV-2 removal were identified. The Centres for Disease Control recommend the consideration of portable HEPA-based air filters; this recommendation applies only to dental facilities where there is deemed to be a high risk of aerosol generation^26^.

This study has limitations, being conducted rapidly in active wards during an ongoing pandemic. The evaluation was conducted in two rooms and there are no data defining the optimal air changes required to remove detectable pathogens with the specified devices. Given the large volume of air within the room and the stability of viruses in the sampling fluid, it was predictable that the amount of SARS-CoV-2 detected via qPCR would be minimal, as evidenced by high C_*T*_ values. Therefore, we cannot categorically state that there was circulating infectious virus. RNA is sufficient to suggest the virus was present and it has been shown that aerosolised virus can remain infectious for >3 hours ^27,28^; additionally, air sampling devices can artefactually reduce the apparent viability of sampled virus. Negative results from the control sampler, and the striking but reversible effect of the air filtration devices, suggest these are not false positive detections and we cannot exclude the risk of airborne infection. Future studies should examine whether air filtration devices, such as those used here, have an impact on healthcare professional and patient focussed outcomes, including measuring infection/exposure as an endpoint, as well as assessing potential harm, such as noise, reduced ambient humidity or impact on delivery of care.

In conclusion, we were able to detect airborne SARS-CoV-2 RNA in a repurposed COVID-19 ‘surge ward’ and found that air filtration can remove SARS-CoV-2 RNA below the limit of qPCR detection. SARS-CoV-2 was infrequently detected in the air of a ‘surge ICU’; however, the device retained its ability to reduce microbial bioaerosols. Our data is highly indicative of aerosolised SARS-CoV-2 circulating in areas that are not classically considered ‘aerosol risk areas’. Furthermore, portable air filtration devices can mitigate the reduced availability of airborne infection isolation facilities when surges of COVID-19 patients overwhelm healthcare resources. The use of such systems may provide additional safety for those that are of high exposure risk to respiratory pathogens such as SARS-CoV-2.

## Data Availability

Data is available on request to the corresponding author.

## References

1. Greenhalgh T, Jimenez JL, Prather KA, Tufekci Z, Fisman D, Schooley R. Ten scientific reasons in support of airborne transmission of SARS-CoV-2. Lancet. 2021;397(10285):1603–1605

2. Chia PY, Coleman KK, Tan YK, et al. Novel Coronavirus Outbreak Research Team. Detection of air and surface contamination by SARS-CoV-2 in hospital rooms of infected patients. Nat Commun. 2020;11(1):2800

3. Zhou J, Otter JA, Price JR, et al. Investigating SARS-CoV-2 surface and air contamination in an acute healthcare setting during the peak of the COVID-19 pandemic in London. Clin Infect Dis. 2020 Jul 8:ciaa905.

4. Nguyen LH, Drew DA, Graham MS, et al. Coronavirus Pandemic Epidemiology Consortium. Risk of COVID-19 among front-line health-care workers and the general community: a prospective cohort study. Lancet Public Health. 2020;5:e475–e483

5. Illingworth C, Hamilton W, Warne B, et al. Superspreaders drive the largest outbreaks of hospital onset COVID-19 infection. OSF Preprints, 15 Feb. 2021. https://doi.org/10.31219/osf.io/wmkn3

6. Sims MD, Maine GN, Childers KL, et al; BLAST COVID-19 Study Group. COVID-19 seropositivity and asymptomatic rates in healthcare workers are associated with job function and masking. Clin Infect Dis. 2020 Nov 5:ciaa1684. Epub ahead of print.

7. Fennelly KP. Particle sizes of infectious aerosols: implications for infection control. Lancet Respir Med. 2020;8:914–924.

8. Meredith LW, Hamilton WL, Warne B, et al. Rapid implementation of SARS-CoV-2 sequencing to investigate cases of health-care associated COVID-19: a prospective genomic surveillance study. Lancet Infect Dis. 2020;20:1263–1271

9. National Institute for Occupational Safety and Health National Institute for Occupational Safety and Health. Hierarchy of controls. Available from https://www.cdc.gov/niosh/topics/hierarchy/default.html (accessed 29/6/21)

10. Morawska L, Allen J, Bahnfleth W, et al. A paradigm shift to combat indoor respiratory infection. Science. 2021;372:689–691

11. UK scientific advisory group for emergencies. Potential application of air cleaning devices and personal decontamination to manage transmission of COVID-19, 4 November 2020 (https://www.gov.uk/government/publications/emg-potential-application-of-air-cleaning-devices-and-personal-decontamination-to-manage-transmission-of-covid-19-4-november-2020)

12. Mousavi ES, Kananizadeh N, Martinello RA, Sherman JD. COVID-19 Outbreak and Hospital Air Quality: A Systematic Review of Evidence on Air Filtration and Recirculation. Environ Sci Technol. 2021;55:4134–4147.

13. Liu DT, Phillips KM, Speth MM, Besser G, Mueller CA, Sedaghat AR. Portable HEPA Purifiers to Eliminate Airborne SARS-CoV-2: A Systematic Review. Otolaryngol Head Neck Surg. 2021:1945998211022636

14. Lindsley WG, Schmechel D, Chen BT. A two-stage cyclone using microcentrifuge tubes for personal bioaerosol sampling. J Environ Monit. 2006;8:1136–42.

15. Killingley B, Greatorex J, Digard P, et al The environmental deposition of influenza virus from patients infected with influenza A(H1N1)pdm09: Implications for infection prevention and control. J Infect Public Health. 2016;9:278–88.

16. Coleman KK, Sigler W V. Airborne Influenza A Virus Exposure in an Elementary School. Sci Rep. 2020 Feb 5;10:1859.

17. Blachere FM, Lindsley WG, Slaven JE, et al. Bioaerosol sampling for the detection of aerosolized influenza virus. Influenza Other Respir Viruses. 2007;1:113–20

18. Blachere FM, Lindsley WG, Pearce TA, et al. Measurement of airborne influenza virus in a hospital emergency department. Clin Infect Dis. 2009;48:438–40.

19. Sridhar S, Forrest S, Kean I, et al. A blueprint for the implementation of a validated approach for the detection of SARS-Cov2 in clinical samples in academic facilities. Wellcome Open Res. 2020;5:110

20. Wilson NM, Marks GB, Eckhardt A, et al. The effect of respiratory activity, non-invasive respiratory support and facemasks on aerosol generation and its relevance to COVID-19. Anaesthesia. 2021 Mar 30. doi: 10.1111/anae.15475. Epub ahead of print.

21. Shields A, Faustini SE, Perez-Toledo M, et al SARS-CoV-2 seroprevalence and asymptomatic viral carriage in healthcare workers: a cross-sectional study. Thorax 2020;75:1089–1094

22. Stockwell RE, Ballard EL, O’Rourke P, Knibbs LD, Morawska L, Bell SC. Indoor hospital air and the impact of ventilation on bioaerosols: a systematic review. J Hosp Infect. 2019;103:175–184.

23. Eckmanns T., Ruden H., Gastmeier P. The influence of high-efficiency particulate air filtration on mortality and fungal infection among highly immunocompromised patients: a systematic review. J Infect Dis. 2006;193:1408–1418

24. Wölfel R, Corman VM, Guggemos W, et al. Virological assessment of hospitalized patients with COVID-2019. Nature. 2020;581:465–469

25. Hamed I, Shaban N, Nassar M, et al. Paired Nasopharyngeal and Deep Lung Testing for Severe Acute Respiratory Syndrome Coronavirus-2 Reveals a Viral Gradient in Critically Ill Patients: A Multicenter Study. Chest. 2021;159:1387–1390.

26. Centers for Disease Control and Prevention [Internet]. Atlanta: The Centers. Interim Infection Prevention and Control Guidance for Dental Settings During the COVID-19 Response. Available from: https://www.cdc.gov/coronavirus/2019-ncov/hcp/dental-settings.html.

27. Fears AC, Klimstra WB, Duprex P, et al. Persistence of Severe Acute Respiratory Syndrome Coronavirus 2 in Aerosol Suspensions. Emerg Infect Dis. 2020 Sep;26(9):2168–71.

28. N van Doremalen N, Bushmaker T, Morris DH, et al. Aerosol and Surface Stability of SARS-CoV-2 as Compared with SARS-CoV-1. N Engl J Med. 2020;382(16):1564–1567.

