## Supplemental methods for "The removal of airborne SARS-CoV-2 and other microbial bioaerosols by air filtration on COVID-19 surge units"

*Air changes in wards*

Both the room in the ‘surge’ ward and ‘surge’ ICU were passively ventilated, without forced air changes.

*Air filtration devices*

The devices used were a AC1500 HEPA14/UV steriliser (Filtrex, Harlow, UK), whilst in the ICU we installed a Medi 10 HEPA13/UV steriliser (Max Vac, Zurich, Switzerland). The filter system has three stage particulate system: a coarse panel pre-filter, a secondary V-flow filter (ePM1=80%), and a HEPA filter, tested to EN1822 standards and >99.99% efficient at removing 0.3-micron particles. The filters are consistently exposed to 253nm UV-C lamps, certified to be 100% effective in removing microbiological agents. The units are certified to supply ISO5-EN ISO 14644 Cleanroom standard air (Class 100 US FED 209E).

*National Institute for Occupational Safety and Health (NIOSH) BC 251 two-stage cyclone aerosol samplers*

Each sampler collects large (>4 μM) particles into a 15 mL centrifuge tube, medium (1–4 μM) particles into a 1.5 mL centrifuge tube, and small (<1 μM) particles in a 37-mm diameter, polytetrafluoroethylene filter with 3-μm pores^S1^. The pump flow rate was set at 3.5 L of air min^−1^, using a flow calibrator and sampling duration set at six hours (collecting a total of 1,260 L/day), following criteria from previous studies demonstrating the capture of airborne viruses for RT-PCR detection^S2-6^.

*Nucleic acid extraction and polymerase chain reactions (PCR)*

To facilitate solubilisation of nucleic acids, tubes were left on a tube rotator overnight at 4°C in lysis buffer containing 4M guanidine thiocyanate and 0.5% β-mercaptoethanol. After overnight solubilisation, all lysis buffer was removed from tubes and the extraction completed as described by Sridhar *et al*^S7^. All samples were eluted in 100 µl nuclease-free water and stored at -80°C until required for qPCR.

*SARS-CoV-2 PCR*

SARS-CoV-2 was detected in samples using the primers and method described previously^S7^ . Briefly, 5 µl of the nucleic acid extract was combined with 20 µl master mix (12.5 µl 2X Luna Universal Probe One-Step reaction mix, 0.5 µl Wu forward and reverse primers (20 pmoles/µl), 0.3 µl Wu FAM-MGB probe (10 pmoles/µl), 0.5 µl MS2 forward and reverse primers (10 pmoles/μl), 0.3 µl MS2 ROX probe (10 pmoles/µl), 1 µl Luna WarmStart RT Enzyme Mix (New England Biolabs, Hitchin, UK) and 3.9 µl nuclease-free water) in a 96-well plate. Reactions were then run on the QuantStudio 7 Flex real-time PCR system (Thermofisher Scientific, Waltham, MA, USA) using the following cycling conditions: 2 minutes at 25°C, 15 minutes at 50°C (reverse transcription), 2 minutes at 90°C and then 45 cycles of 3 seconds at 95°C followed by 30 seconds at 60°C.

*Bioaerosol high-throughput qPCR*

Other pathogens were detected using a BioMark HD qPCR system (Fluidigm, Cambridge, UK). To prepare individual 10X assays for the BioMark HD qPCR, 2.5 µl of each forward and reverse primer pair (100 µM), was combined with 25 µl of 2X Assay Loading Reagent and 22.5 µl of TE buffer to a final primer concentration of 500nM. Microbial targets are listed in Table S1. Pooled assays for pre-amplification were produced by combining 1µl of all primer pairs and diluting to a final volume of 200 µl in TE buffer (Invitrogen, Thermofisher Scientific) to give a final primer concentration of 500 nM. Stock solutions of the pooled and individual assay mixtures were stored at −20°C.

4µl of nucleic acid extract was first reverse-transcribed using Fluidigm Reverse Transcriptase as per manufacturer instructions. Pre-amplification of cDNA was then performed to minimise sampling bias, using the Fluidigm PreAmp Master Mix Kit. 1.25 µl of reverse transcribed samples were then combined with 2.5 µl 2X PreAmp Master Mix, 0.5 µl pooled primers (500nM), 0.75 µl and nuclease-free water. Reactions were then run using cycling conditions of 95 °C for 10 minutes, followed by 17 cycles of 95 °C for 15 seconds and 60 °C for 2 minutes, and a final hold at 4 °C. Finally, samples underwent exonuclease I (Exo-I) (NEB) treatment to degrade any remaining single stranded DNA in accordance with manufacturer’s instructions, before dilution 1:5 with TE buffer.

Samples were prepared for IFC (integrated fluidics circuit) loading as per manufacturer’s instructions, with 2.5 µl of 2× SsoFast™ EvaGreen^®^ Supermix Low ROX (BioRad, Watford, UK) and 0.25 µl of 20× DNA Binding Dye Sample Loading Reagent combined with 2.25 µl of the Exo-I treated samples. 5 µl of each assay mix (see above) and sample mix was loaded into the suitable IFC inlets and the IFC was loaded using the Fluidigm Juno. Once complete, the IFC was moved to the BioMark HD for qPCR using the pre-programmed thermal protocol: GE Fast 96x96 PCR+Melt v2.pcl.

Preliminary thresholding of the amplification data was completed using the Fluidigm Real-Time PCR Analysis Software, before raw data was exported to R (RStudio, Boston, USA) to apply manually defined melting curve peak thresholds. Positive samples were determined to be those with Ct values <= 23 and with melt curves within the previously determined range for that assay target.

**Supplemental Table 1.** Bacterial, fungal, and viral targets which formed the targets of the microbial bioaerosol high-thoughput qPCR*.

| ***Bacteria*** | **Mycobacteria** | **Atypical bacteria** | **Fungi** | **Viruses** |
| --- | --- | --- | --- | --- |
| *Acinetobacter baumannii* | *Mycobacterium tuberculosis* | *Chlamydia pneumoniae* | *Aspergillus fumigatus* | Adenovirus |
| *Bordetella pertussis* | *Mycobacterium* spp | *Chlamydia psittaci* | *Aspergillus* spp | Bocavirus |
| *Bordetella parapertussis* |  | *Coxiella burnetii* | *Candida* spp | HCoV 229E |
| *Citrobacter* spp |  | *Legionella pneumophila* | Fungal ribosomal 18S | HCoV NL63 |
| *Corynebacterium diphtheriae* |  | *Legionella* spp |  | HCoV OC43 |
| *Escherichia coli* |  | *Mycoplasma pneumoniae* |  | HCoV HKU1 |
| *Enterococcus faecium* |  | *Leptospira* spp |  | Cytomegalovirus |
| *Enterococcus faecalis* |  |  |  | Epstein-Barr virus |
| *Enterococcus* sp |  |  |  | Enterovirus |
| *Elizabethkingia meningoseptica* |  |  |  | Herpes Simplex virus |
| *Haemophilus influenzae* |  |  |  | Influenza A virus |
| *Klebsiella pneumoniae* |  |  |  | Influenza B virus |
| *Moraxella catarrhalis* |  |  |  | Human Metapneumovirus |
| *Morganella morganii* |  |  |  | Measles morbillivirus |
| *Neisseria meningitidis* |  |  |  | Mumps virus |
| *Proteus mirabilis* |  |  |  | Parainfluenza |
| *Pseudomonas aeruginosa* |  |  |  | Parechovirus |
| *Serratia marcescens* |  |  |  | Rhinovirus |
| *Staphylococcus aureus* |  |  |  | Respiratory syncytial virus |
| *Staphylococcus epidermidis* |  |  |  | Rubella virus |
| Coagulase negative staphylococci |  |  |  | SARS-CoV-2 |
| *Stenotrophomonas maltophilia* |  |  |  | Varicella zoster virus |
| *Streptococcus pneumoniae* |  |  |  |  |
| *Streptococcus pyogenes* |  |  |  |  |

*Species were selected for their known respiratory pathogenicity or frequency as agents of hospital-acquired infection. HCoV human corona virus, SARS-CoV-2 severe acute respiratory syndrome coronavirus 2. Loading control was with bacteriophage MS2. (Primer sequences available on request**)**

**Supplemental references**

S1.	Lindsley WG, Schmechel D, Chen BT. A two-stage cyclone using microcentrifuge tubes for personal bioaerosol sampling. *J Environ Monit.* 2006 ;8:1136-42.

S2.	Chia PY, Coleman KK, Tan YK, et al. Novel Coronavirus Outbreak Research Team. Detection of air and surface contamination by SARS-CoV-2 in hospital rooms of infected patients. *Nat Commun*. 2020;11(1):2800.

S3.	Killingley B, Greatorex J, Digard P, et al The environmental deposition of influenza virus from patients infected with influenza A(H1N1)pdm09: Implications for infection prevention and control. *J Infect Public Health*. 2016;9:278-88.

S4.	Coleman KK, Sigler W V. Airborne Influenza A Virus Exposure in an Elementary School. *Sci Rep*. 2020 Feb 5;10:1859.

S5.	Blachere FM, Lindsley WG, Slaven JE, et al. Bioaerosol sampling for the detection of aerosolized influenza virus. *Influenza Other Respir Viruses.* 2007;1:113-20.

S6.	Blachere FM, Lindsley WG, Pearce TA, et al. Measurement of airborne influenza virus in a hospital emergency department. *Clin Infect Dis*. 2009;48:438-40.

S7.	Sridhar S, Forrest S, Kean I, et al. A blueprint for the implementation of a validated approach for the detection of SARS-Cov2 in clinical samples in academic facilities. *Wellcome Open Res*. 2020;5:110
